# Psychometric properties and factor invariance for the General Health Questionnaire (GHQ-28): study in Peruvian population exposed to the COVID-19 pandemic

**DOI:** 10.1101/2020.11.10.20229435

**Authors:** Rita J. Ames-Guerrero, Victoria A. Barreda-Parra, Julio C. Huamani-Cahua

## Abstract

Large-scale epidemics are known to significantly disrupt the mental health and perceived well-being of most populations. In spite of the wide range of screening tools, there are not many reliable and widespread tools for the identification of psychological symptoms in non-clinical populations during a health crisis.

**Objective:** The aim of this study was to conduct a psychometric analysis of the Goldberg’s GHQ-28 (1) through a sample of Peruvian adults using confirmatory factor analysis.

**Materials and methods:** 434 individuals have been examined, studying the goodness and structure of the Goldberg GHQ-28 questionnaire.

**Result:** We found high reliability indicating optimal values (α by Cronbach = .829), also there are four correlated factors that show strict invariance among the 28 items. Confirmatory factor analysis (CFA) and exploratory factor analysis (EFA) were used to examine the structure of the subscales. There are high levels of anxiety (X=1.01) and social dysfunction (X=1.21) in the assessed sampling.

**Conclusion:** The factorial structure obtained in this study is similar to that originally described by the researchers involved in the original questionnaire. It is concluded that GHQ-28 is suitable to explore prevalence of psychopathologies in emergency contexts and social isolation for general non-psychiatric population.

## 1. Introduction

Health, understood as a “sense of general well-being and not just the absence of disease”, as mentioned by the WHO, is an essential condition for the continuity and integral development of individuals. Maintaining adequate overall health in crisis contexts then becomes a growing challenge for health managers on a global scale. In this context, it should be noted that while there are sustained efforts to implement public protection strategies to improve COVID-related infections,19, reports of prevalence of mental illness due to emotional instability have been increasing throughout the period of the first coronavirus outbreak, this exposes the population to the development of psychopathologies and states of deterioration of mental health (3-5)

Previous studies emphasize the importance of identifying in early stages the psychological symptoms of nonpsychiatric population in order to reduce the probability of developing mental disorders and the chronology of clinical symptoms (6,7). However, despite the extensive list of psychological health screening instruments, there are some diagnostic accuracy problems (8). Within the considerations to choose one instrument of psychological symptom screening over another, For example, criteria of scientific rigor are found to ensure that the instrument has the reliability and validity necessary to optimally integrate the diagnosis and treatment, in addition to containing criteria for adaptation of the content (9).

Accurate diagnosis in mental health is critical to guiding treatment programs (6). While the standard diagnostic guidelines “gold standard” for psychiatric evaluation are the most appropriate, the psychological screenings provide rapid and low-cost diagnostic support for the identification of cases. It is worth mentioning that the use of the General Health Questionnaire (GHQ-28) has been extended to a wide variety of diseases and clinical diagnoses as a self-perceived health evaluation tool. This tool was previously used to identify psychosocial problems related to stressful life events in primary care services, such as in the treatment of chronic diseases whose data on individual variables (age, sex, activity, socioeconomic and cultural level) make it possible to determine the prevalence of health indicators.

Although there are multiple previous validations on the psychometric properties of GHQ-28 with psychiatric patients and/or with physical disease, few studies base their exploration on normal population. In the literature, we found validation studies of the GHQ-28 questionnaire in North America with patients diagnosed with physical deterioration (10); endocrinological diseases in samples from patients with drug addiction (11) as opioid dependence (12). Additionally, we found studies in military samples for example: Farhood et all validated Arabic versions in war-exposed civilians (13), as well as predictive validity studies of GHQ-28 in patients with substance use disorders in therapeutic communities (14) in Spanish-speaking countries.

Previous studies have shown the use of confirmatory and exploratory analysis factors to study the psychometric properties of the questionnaire. In particular, we found a reference study described by Prady et all., who analyses multi-ethnic samples of pregnant women using confirmatory and exploratory analysis for a longitudinal study (15); Similarly, we found confirmatory analysis studies of GHQ-28 with non-clinical samples in adults of the armed forces in South Africa (16) and population with coronary heart disease in Norway, basing the analysis on configural invariances, metrics and scalars for an experimental study design (17).

The clinical evaluation process applied in various populations entails challenges related to the translation and adaptation of the instrument. One relevant aspect to consider are the ethnic and semantic differences in content that might have an “unexpected” response in the results (15). The rapid identification of emotional problems in a single instrument makes it necessary to validate the Goldberg general health questionnaire (GHQ-28). Therefore, it is conceivable that due to the inherent complexities of nature and cultural differences it is appropriate to validate the general health instrument in a non-discretionary population under conditions of global emergency.

## 2. Materials and Methods

### 2.1 Population and sample

An observational, descriptive, cross-sectional study was carried out to analyze psychological health in the general population. During the Covid-19 pandemic, anonymous online questionnaires were administered to the general population in the March-April period. The study population was made up exclusively of people (18 years old) able to give their informed consent through a convenient snowball sampling process. Participants were required to reside in any department of Peru. No monetary compensation was granted for completing the questionnaire. The study protocol was approved by the ethics committee of the Catholic University of Santa María (ref. no 167-2020).

The sample is made up by general population (n=434) where 61.3% are women and 38.7% men. Participants were selected through the convenience method and snowball sampling, during the pandemic by COVID-19 in the period April-June 2020. The recruitment criteria were limited to individuals over 18 years of age residing in any department of Peru with access to smartphones to fill in anonymous online questionnaires, corresponding with an instrumental-type study (Montero and León, 2007).

Regarding sociodemographic data from the sample, 14.7% have secondary education, 40.1% have university studies, 26.7% have a bachelor’s degree and 18.4% have postgraduate studies, The instrument also indicates that 40.8% have psychological disorder and 59.2% are not psychological cases. Table 1 shows the characteristics of the study sample.

**Table 1.**
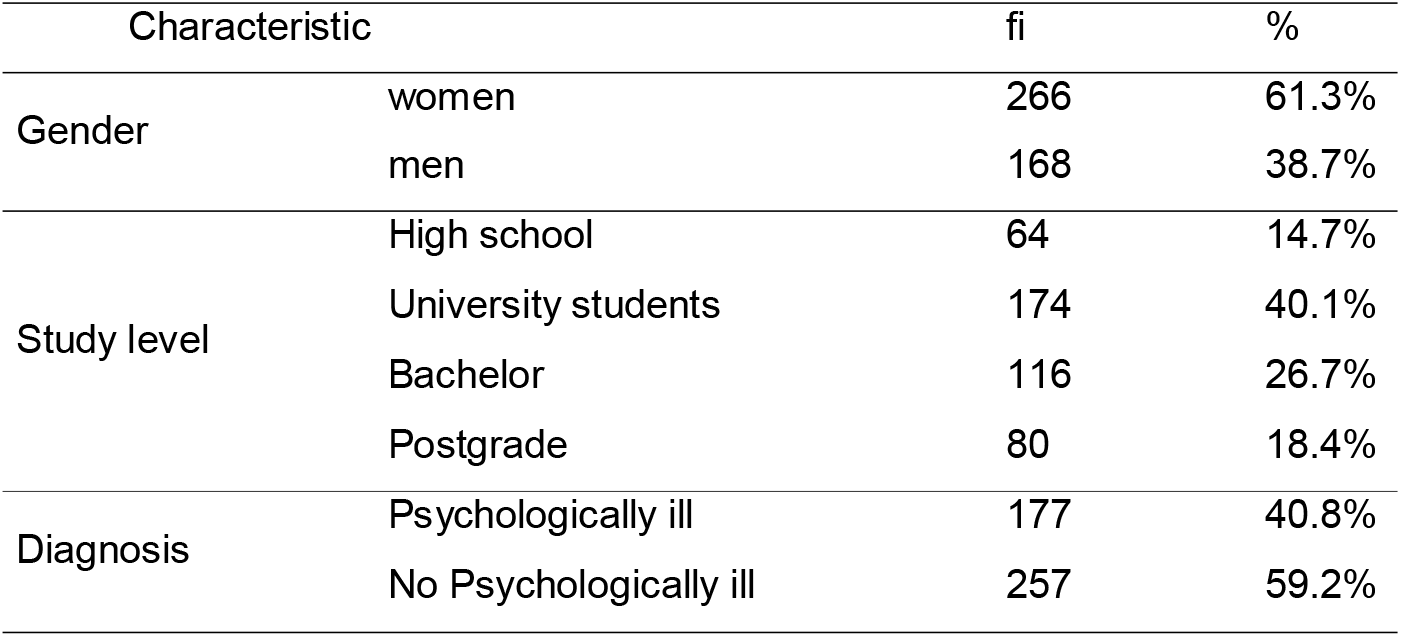
Sampling characteristic

| Characteristic |  | fi | % |
| --- | --- | --- | --- |
| Gender | women | 266 | 61.3% |
|  | men | 168 | 38.7% |
| Study level | High school | 64 | 14.7% |
|  | University students | 174 | 40.1% |
|  | Bachelor | 116 | 26.7% |
|  | Postgrade | 80 | 18.4% |
| Diagnosis | Psychologically ill | 177 | 40.8% |
|  | No Psychologically ill | 257 | 59.2% |

Fully filled instruments were considered valid and the exclusion criteria were under 18 years of age, and those who scored high in bias for social desirability. Prior to the beginning of the study, approval was obtained from the local ethics committee of the Catholic University of Santa Maria, in addition to the written consent of each participant.

### 2.2 Instrument

#### The General Health Questionnaire (GHQ-28)

GHQ-28 is a self-administered general health questionnaire for the detection of mental disorders, which contains 28 itemes. Participants were asked about the symptoms and/or discomfort they had experienced recently (in recent weeks) during the coronavirus pandemic. Each item is graded on a 4-point scale to identify the severity of symptoms between 0 and 3 (“Not at all”, “No more than usual”, “More than usual” and “Much more than usual”). This version contains 4 subscales: somatic symptoms, anxiety, insomnia, social dysfunction, and severe depression (1).

Reliability was calculated with the Cronbach Alpha coefficient and the Omega coefficient, finding optimal values (ω = .829 and ω = .911) (Campo-Arias, & Oviedo, 2008). The values for each factor were: somatic symptoms, ω = 0.829 (95% CI = 0.802-0.852); anxiety/insomnia, ω = 0.911 (95% CI = 0.895-0.922), social dysfunction, ω = 0.800 (95% CI = 0.766-0.824); and severe depression factor, ω = 0.900 (95% CI = 0.875-0.906). The normative data for the general population evaluated indicates a cutoff point ≥ 5 (12).

## 3. Procedure

The general health questionnaire (GHQ-28) was converted to a virtual format to facilitate filling and reduce the risk of infection of researchers. Those who decided to fill in the tool, received information to participate in the study, for this it was requested their informed consent where they were explained the objective of the investigation and the implicit risks.

## 4. Statistical analysis

To perform the psychometric analysis of the Goldberg questionnaire (GHQ-28), the descriptive data (mean, standard deviation, asymmetry, kurtosis and item-test correlation) we used the software JASP, considering a threshold of ± 2 to identify asymmetric values from normal values (17).

Subsequently, the internal structure of the instrument was evaluated through the Confirmatory Factor Analysis (CFA) corresponding to the theoretical model proposed by the author, which indicated four factors and 28 items. This CFA were performed throughout RStudio® software using psych package, lavaan y package for complex survey analysis of structural equation models (18), Tools for Structural Equation Modeling (19) WLSMV estimator (minimum weighted squares with adjusted mean and variance), this estimator presents robustness in the results in situations of nonnormality and categorical nature of the variables. The Comparative Adjustment Index (CAI) was also considered ≥ .90 (17), and the Tucker-Lewis index (TLI), mean quadratic standardized residual root (SRMR) and the approximation mean quadratic error (RMSEA) with values ≤.80 (20).

We performed a multigroup CFA, from that, four types of invariance were used: configural, weak, strong and strict (restricting factor loads, intercepts and variances of errors), the criteria used were the difference in the square Chi, as well as the comparative adjustment indices proposed by Millsap et all., (21) values lower than .010 in the CIF and of .015 in the RMSEA are considered indicators of equivalence for the models. Finally, the reliability of the internal consistency was evaluated with the Cronbach and Omega Alpha coefficient of McDonald (ω) (22), as well as the average extracted variance (AEV), whose value exceeds 0.5 provides evidence of convergent internal validity (23).

## 5. Results

The confirmatory Factor Analysis and exploratory analysis allowed to assess the structure of the (GHQ-28) sub-scales in Peruvian population not psychiatric. In Table 2, the quantitative analysis is shown, we found that global averages varied for the subscales of anxiety/insomnia and social dysfunction, whose means are high and are between (M= 1.01; DE = 0.885) and (M= 1.21; DE = 0.942) for items (1, 5, 8, 9, 10, 11, 15, 16, 17, 18, 20, 21).

**Table 2.**
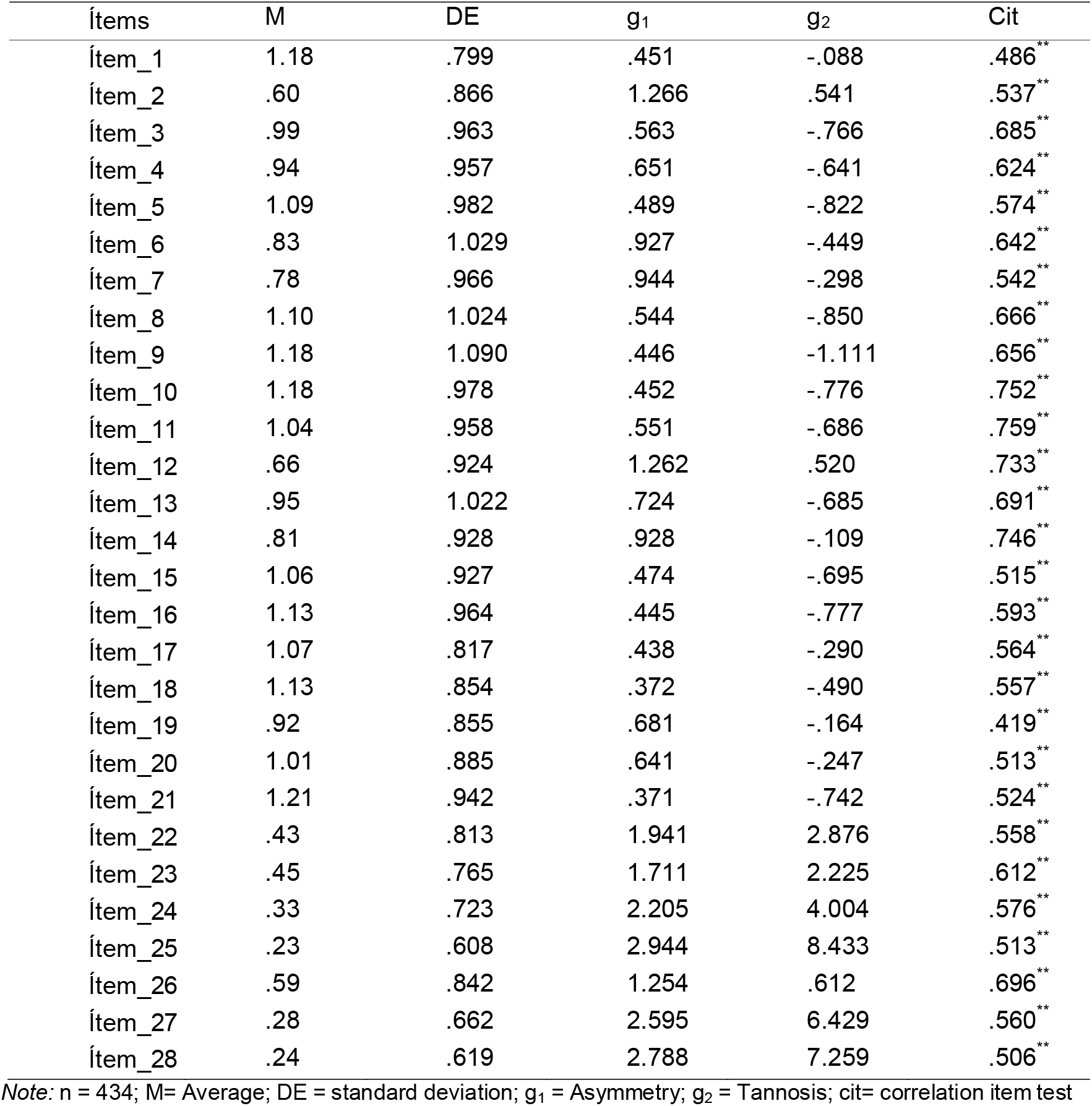
Item analysis

And the other group, formed by the items (2, 3, 4, 6, 7, 12, 13, 14, 19, 22, 23) of the subscales of somatic symptoms and severe depression present low averages (M= 0.23; DE = 0.608) and (M= 0.99; DE = 0.963), with asymmetry values less than 2 (17) indicating slight deviations from normality. It is observed that the correlations between the elements are not greater than 0.90 (24).

We found an appropriate fit, from the four correlated GHQ-28 factor model, of 7 items per factor (χ^2^ = 1179.306, gl = 344, χ^2^/gl = 3.42; IFC = .927; TLI = .919; RMSEA = .075 [90%CI: .07, .08]; SRMR = .07). Table 3 shows the goodness-of-fit index of GHQ-28.

**Table 3.**
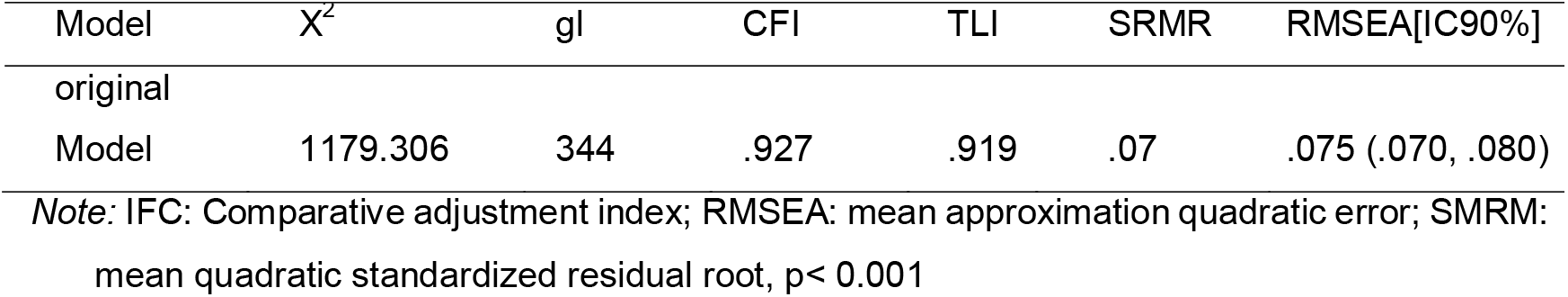
GHQ-28 goodness-of-fit index

Table 4 shows the standardized factor loads that confirm the four-factor model proposed by the author of GHQ-28, with appropriate values λ > .581 (≥ .5) (except ítem 19, λ = .469) (Johnson & Stevens, 2001). Additionally, we observed that the correlation between the factor valuing somatic symptoms and anxiety/insomnia were high (> 0.75), likewise, the correlations between the variables do not show multicollinearity.

**Tabla 4.**
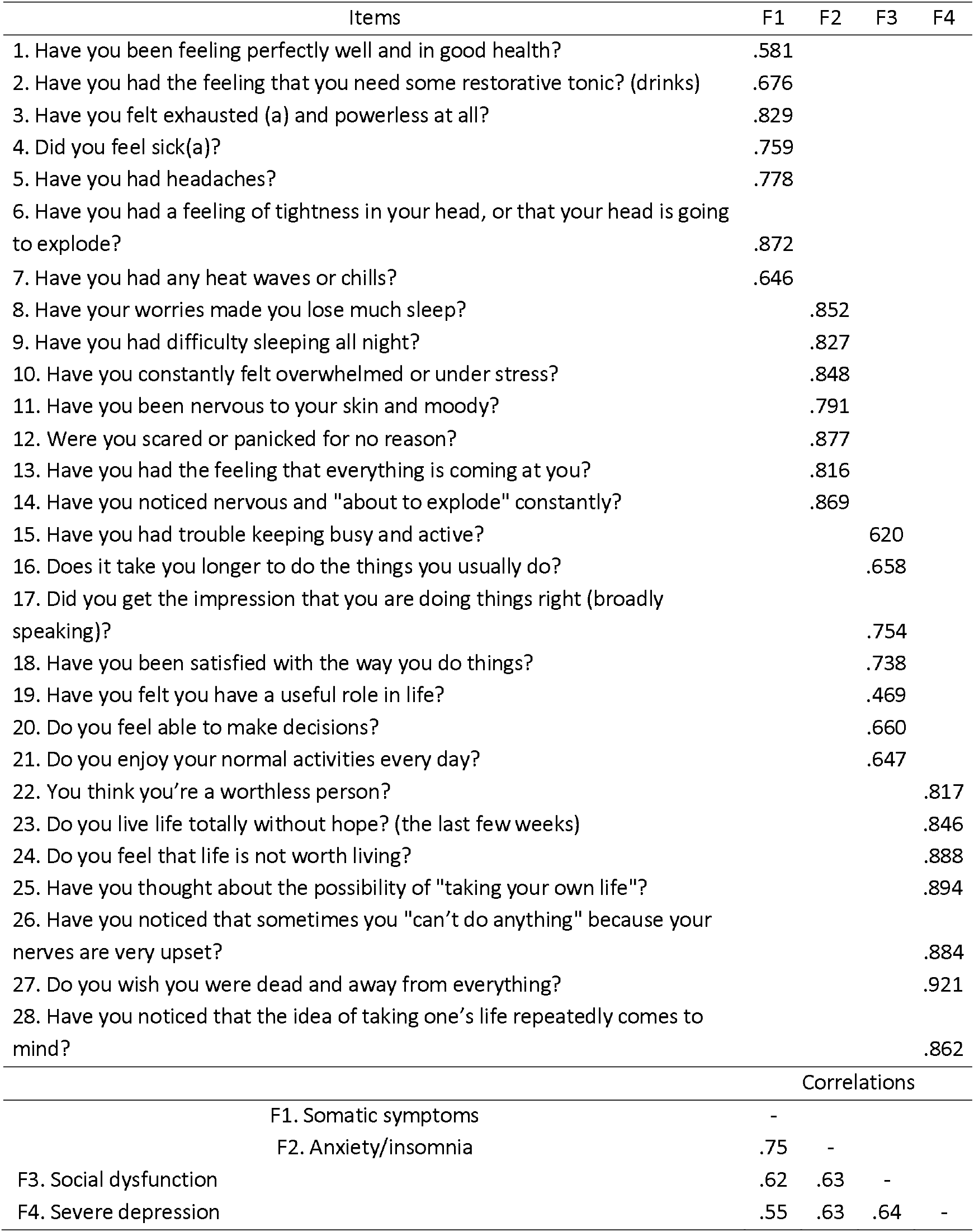
Factorial loads of the AFC standardized solution for the final model

In Table 5, the invariance of the four correlated factors from the CFA (confirmatory factor analysis) is shown. We found strict invariance, it is to be noted that the factorial loads are similar in the group of participants with diagnosis of psychological disorder and non-psychological cases, as well as in the group according to level of studies. However, according to gender (men and women), invariance is evidenced, where the configural model (baseline) presents adequate fit indices X2 (gl) = 595.11 (688), CFI = 0.90, RMSEA = 0.042, with reference to the model metric (weak invariance), scalar (strong invariance) and strict invariance.

**Table 5.**
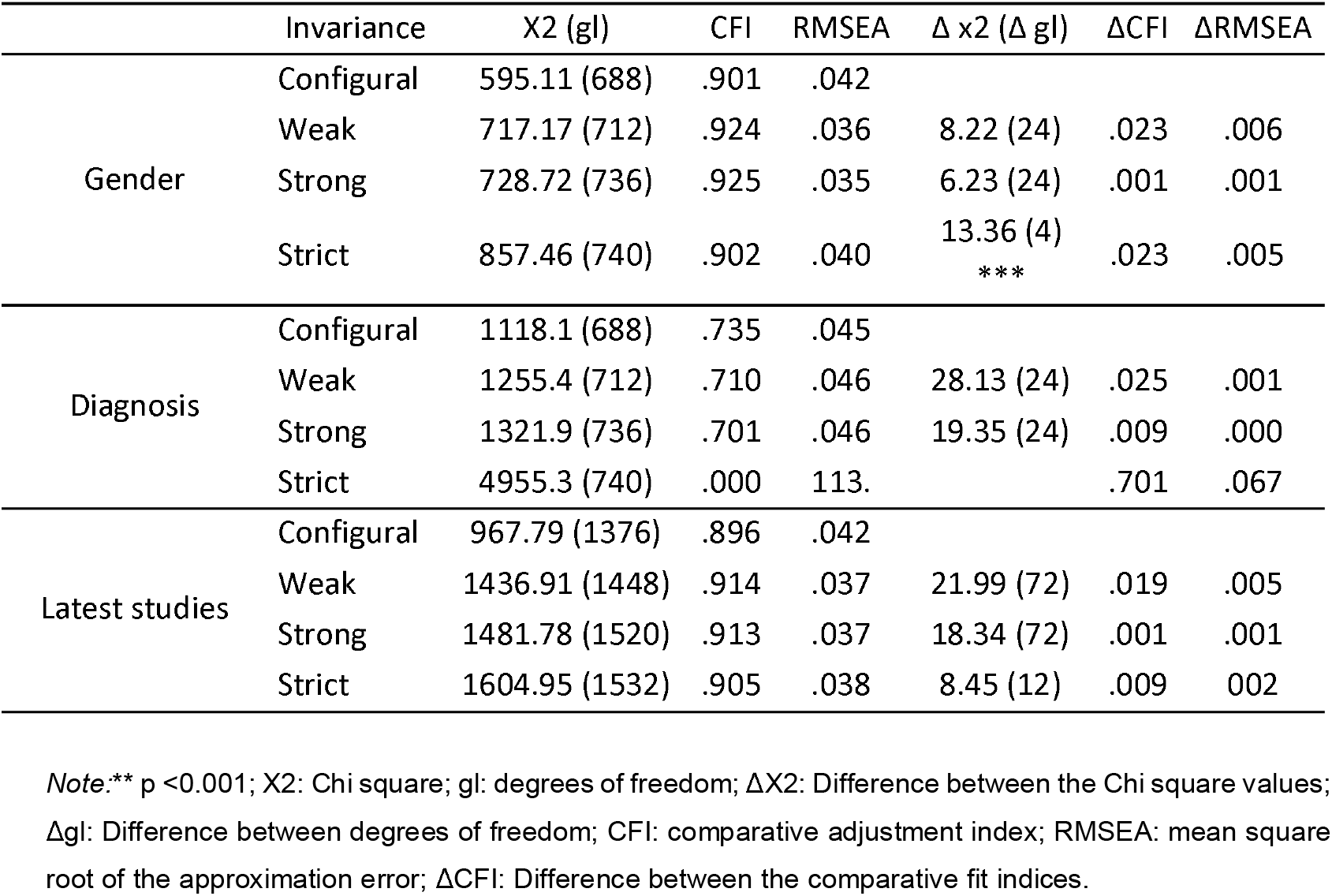
Measurement invariance for the GHQ-28 four-factor model, according to gender, diagnosis and latest studies performed

**Table 6.**
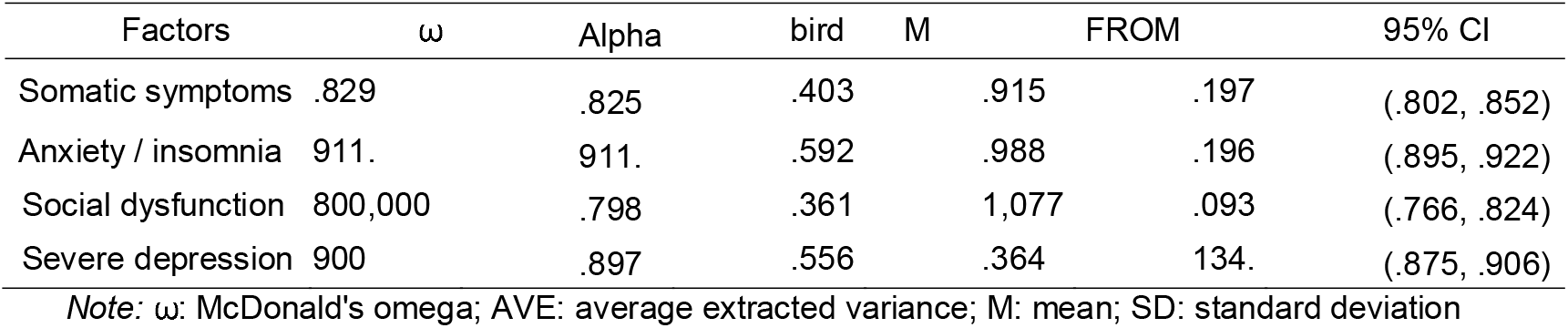
Reliability coefficients and descriptive measures of the GHQ-28 four-factor model.

| Factors | $\omega$ | Alpha | bird | M | FROM | 95% CI |
| --- | --- | --- | --- | --- | --- | --- |
| Somatic symptoms | .829 | .825 | .403 | .915 | .197 | (.802, .852) |
| Anxiety / insomnia | .911 | .911 | .592 | .988 | .196 | (.895, .922) |
| Social dysfunction | .800,000 | .798 | .361 | 1,077 | .093 | (.766, .824) |
| Severe depression | .900 | .897 | .556 | .364 | .134 | (.875, .906) |

*Note:* $\omega$ : McDonald's omega; AVE: average extracted variance; M: mean; SD: standard deviation

The factor loadings between men and women are equal. There are no statistically significant differences (p> .05) and (ΔCFI ≤ .01), (ΔRMSEA ≤ .015) when comparing with the base model (configural), the values found are below the cut-off points established with respect to the metric invariance.

Finally, the strict invariance is observed (restrictions on factor loads, intercepts and residuals) that indicates variance in the group of men and women, with statistically significant differences (p <.05) and (ΔCFI ≤ .01), the values found were (p = 0.001), (ΔCFI ≤ .023), to verify these differences, the Student’s t test and effect sizes were used, using the *d* of Cohen (1992), the referential values are, d = 0.20 (small), d = 0.50 (medium) and d = 0.80 (large), finding statistically significant differences and median TE in two factors, somatic symptoms (p <0.01; d = 41) and anxiety / insomnia (p <0.01; d = 35), the other two factors (social dysfunction and severe depression) did not show differences.

In addition, the mean of the extracted variance (AVE> 0.5) was included, indicating convergent validity, where the factors (somatic symptoms and social dysfunction) do not meet this criterion.

## 6. Discussion

In this study, the psychometric properties of the Goldberg general health questionnaire for non-psychiatric population were determined in the Spanish version for Peruvian adults using confirmatory factor analysis. This analysis would indicate that the internal factorial structure of the instrument according to the four-factor model yielded an adequate fit.

Consistent with our findings, the factor analysis of previous studies also reported the construct validity of four factors with samples from different patients. The various versions found confirm that the four-factor model, such as: Malaysian version with patients with drug addiction, diabetes and normal population (9), in the Spanish version to patients with fibromyalgia (18), in the Norwegian version to patients who suffered a cerebrovascular accident, with some differences in the factor structure compared to the original version by Goldberg and Hillier (17), also in the Spanish version with patients diagnosed of opiate dependence. However, despite the fact that the 4-factor model is significantly greater than the 3-factor model, as proposed by Goldberg and Hillier(12), the proper fit of the original instrument was not achieved.

It should be noted that the studies conducted on non-clinical samples of different ethnicities did not obtain a good statistical fit. For example, the three-factor structure fit better than the four-factor scale in a South African black sample of military employees.(16), and the five-factor structure had a better fit in a sample of maternal women of diverse ethnicity and languages in the United Kingdom (fifteen). However, in recent studies with clinical and non-clinical samples, psychological health has been evaluated with the four-factor GHQ-28, according to the original version of Goldberg 1972 and Goldberg and Hillier 1979(19–21).

It is necessary to point out that with regard to invariance, this is the first study to examine the measurement invariance of the GHQ-28 of four factors in the three groups formed: a) gender, b) diagnosis and c) latest studies. Strict invariance is accepted in the group of participants with a diagnosis of psychological disorder and non-psychological cases and in the group of recent studies carried out, that is, it is accepted that factor loadings, factor weights and intercepts are similar in each group. Additionally, strict variance is found according to gender, finding statistically significant differences (median effect size) in two factors, somatic symptoms and anxiety / insomnia, however the other two factors: social dysfunction and severe depression show no differences.(22) who reports that the most stable factors were social dysfunction and depression, indicating a great invariance in terms of their factorial composition in the total sample (58% women) of primary health care patients.

Another important finding is the high internal consistency of the anxiety / insomnia subscale (α = .798 and α .911 and ω = .829 and ω = .911 and α = .79) consistent with previous studies, where all the scales correlate positively and significantly with each other and with the total scale. In the Arabic version(13) internal consistency was .91 and .80 in civilian population living in war zones, in the Malay version (9) it was from .859 to .915 in the normal population and with patients. Acceptable values between .70 and .83 were reported in a sample of military in South Africa (16) and values between .719 and .881 in the Norwegian version with patients who suffered a stroke (17).

In this study, GHQ-28 was applied during the COVID-19 coronavirus pandemic, just as it was applied in other countries for the detection of psychological problems in the general population. In the sample of Peruvian participants, 40.8% reported psychological problems and obtained higher means in the subscales of anxiety and insomnia and social dysfunction, 61.3% of the total population were women. Being a woman was associated with a higher risk of suffering from anxiety and insomnia and somatic symptoms(3). These results are similar to those reported by a study reported in China (21), 42.65% had a high prevalence of psychological problems (GHQ-28 ≥ 5), 48.3% on the subscale of depression, 22.6% in the anxiety subscale and 19.4% in a combination of both, with a higher risk in the group of 18 and 39 years, students and technical and professional employees, 56.09% were men. Higher percentages were found in Poland with mixed samples and higher participation of women. In the sample of medical and non-medical professionals, psychological problems were moderate (total score GHQ-28> 24) in 60.8% of the medical group vs 48% of the non-medical group, in two subscales, somatic symptoms and anxiety and insomnia, with a higher risk for women, 74.4% of the non-medical group were women (twenty). In another longitudinal study in Poland with an alcohol consuming population, compared with the group that did not consume alcohol (27.8%), no differences were found in the somatic and anxiety/ insomnia subscale, with a higher risk of suffering from depressive symptoms and worse mental health in participants who consumed more alcohol than before, during the pandemic, 78% of the total population were women (19).

In summary, except in China with a relatively higher percentage of male participants (56.09%), depressive symptoms are not significant in the normal population, in all samples with a higher participation of women, symptoms of anxiety and insomnia were reported as a consequence of the pandemic. In another study conducted before the pandemics, there was a higher participation of men (83.6%) with a higher average (12,42) in the social dysfunction subscale (16). These studies confirm that gender is an elemental factor associated with anxiety, insomnia and somatic symptoms.

In the review of the literature, we found no evidence of the validation of the GHQ-28 in the Latin American population, so we consider that this study is a contribution to the theoretical and empirical knowledge of the instrument. Unlike other questionnaires, this study examines the invariance of four factors by gender, diagnosis (psychological disorder and no psychological case) and educational level (secondary, university studies, undergraduate and graduate).

These findings represent a significant input to suggest the use of the GHQ-28 in the identification of the general psychological health status, which, according to the analysis carried out, must take into account the gender, age and educational level approach that could serve as a predictive instrument in contexts of global uncertainty.

Although the results are not conclusive, the evaluation of anxiety with other instruments (different from the GHQ-28) concluded similar patterns, finding differences by gender, with a higher percentage of women reporting anxiety symptoms above the cut-off point (2. 3) and others who did not report gender differences in anxiety to coronavirus (24). Our findings were consistent with Huang’s Asian study.(26) who suggests a high correlation between anxiety, somatization and the coexistence of insomnia symptoms in the general population (> 0.75).

Among the possible limitations, the validation of the GHQ-28 cannot be considered representative of the entire Peruvian population, given the non-probabilistic nature of the sample, the non-inclusion of participants of various ethnicities and languages; Future studies could focus on the population with low economic resources, in marginal urban areas, with low academic achievements, and without connection to wireless networks. Even though, it is not a random and large population-based study, our sample represents Peruvians from various departments of Peru.

## 7. Conclusion

The factorial structure of the GHQ-28 confirms the four-factor model, especially suitable for working in the Peruvian context with an adult population. An important strength of the study is that the instrument presents factorial invariance and can be useful for psychological evaluation, it has also been shown that it presents adequate reliability for this population.

Therefore, the questionnaire is useful for making diagnostic assumptions in people seeking help and who may require psychotherapeutic support.

## Supporting information

www.table.net

## Data Availability

data is available

## Contribution of the authors

RAG. conceived and designed the study. RAG, JHC, ABP collected the data, JHC analyzed and interpreted the data. All authors participate in the writing and approval of the final version of the article.

## Financing sources

This study had no funding

## Ethical approval and consent to participate

Research approaval was obtained from the ethics committee of the Catholic University of Santa María (ref. No. 167-2020), the participants gave consent before being included in the study.

## Acknowledgment

This study was part of a series of investigations on mental health. We thank the teachers and students who were involved in the development of this study. The authors declare no conflict of interest.

