## Supplementary material for "Psychometric properties and factor invariance for the General Health Questionnaire (GHQ-28): study in Peruvian population exposed to the COVID-19 pandemic": www.table.net

Table 2

*Item analysis*

| Ítems | M | DE | $g_1$ | $g_2$ | Cit |
|---|---|---|---|---|---|
| Ítem_1 | 1.18 | .799 | .451 | -.088 | .486** |
| Ítem_2 | .60 | .866 | 1.266 | .541 | .537** |
| Ítem_3 | .99 | .963 | .563 | -.766 | .685** |
| Ítem_4 | .94 | .957 | .651 | -.641 | .624** |
| Ítem_5 | 1.09 | .982 | .489 | -.822 | .574** |
| Ítem_6 | .83 | 1.029 | .927 | -.449 | .642** |
| Ítem_7 | .78 | .966 | .944 | -.298 | .542** |
| Ítem_8 | 1.10 | 1.024 | .544 | -.850 | .666** |
| Ítem_9 | 1.18 | 1.090 | .446 | -1.111 | .656** |
| Ítem_10 | 1.18 | .978 | .452 | -.776 | .752** |
| Ítem_11 | 1.04 | .958 | .551 | -.686 | .759** |
| Ítem_12 | .66 | .924 | 1.262 | .520 | .733** |
| Ítem_13 | .95 | 1.022 | .724 | -.685 | .691** |
| Ítem_14 | .81 | .928 | .928 | -.109 | .746** |
| Ítem_15 | 1.06 | .927 | .474 | -.695 | .515** |
| Ítem_16 | 1.13 | .964 | .445 | -.777 | .593** |
| Ítem_17 | 1.07 | .817 | .438 | -.290 | .564** |
| Ítem_18 | 1.13 | .854 | .372 | -.490 | .557** |
| Ítem_19 | .92 | .855 | .681 | -.164 | .419** |
| Ítem_20 | 1.01 | .885 | .641 | -.247 | .513** |
| Ítem_21 | 1.21 | .942 | .371 | -.742 | .524** |
| Ítem_22 | .43 | .813 | 1.941 | 2.876 | .558** |
| Ítem_23 | .45 | .765 | 1.711 | 2.225 | .612** |
| Ítem_24 | .33 | .723 | 2.205 | 4.004 | .576** |
| Ítem_25 | .23 | .608 | 2.944 | 8.433 | .513** |
| Ítem_26 | .59 | .842 | 1.254 | .612 | .696** |
| Ítem_27 | .28 | .662 | 2.595 | 6.429 | .560** |
| Ítem_28 | .24 | .619 | 2.788 | 7.259 | .506** |

*Note:* n = 434; M= Average; DE = standard deviation; $g_1$ = Asymmetry; $g_2$ = Tannosis; cit= correlation item test

**Table 3**

***GHQ-28 goodness-of-fit index***

| Model | $X^2$ | gl | CFI | TLI | SRMR | RMSEA[IC90%] |
|---|---|---|---|---|---|---|
| original Model | 1179.306 | 344 | .927 | .919 | .07 | .075 (.070, .080) |

*Note:* IFC: Comparative adjustment index; RMSEA: mean approximation quadratic error; SMRM: mean quadratic standardized residual root, p< 0.001

**Tabla 4**

***Factorial loads of the AFC standardized solution for the final model***

| Items | F1 | F2 | F3 | F4 |
|---|---|---|---|---|
| 1. Have you been feeling perfectly well and in good health? | .581 | | | |
| 2. Have you had the feeling that you need some restorative tonic? (drinks) | .676 | | | |
| 3. Have you felt exhausted (a) and powerless at all? | .829 | | | |
| 4. Did you feel sick(a)? | .759 | | | |
| 5. Have you had headaches? | .778 | | | |
| 6. Have you had a feeling of tightness in your head, or that your head is going to explode? | .872 | | | |
| 7. Have you had any heat waves or chills? | .646 | | | |
| 8. Have your worries made you lose much sleep? | | .852 | | |
| 9. Have you had difficulty sleeping all night? | | .827 | | |
| 10. Have you constantly felt overwhelmed or under stress? | | .848 | | |
| 11. Have you been nervous to your skin and moody? | | .791 | | |
| 12. Were you scared or panicked for no reason? | | .877 | | |
| 13. Have you had the feeling that everything is coming at you? | | .816 | | |
| 14. Have you noticed nervous and "about to explode" constantly? | | .869 | | |
| 15. Have you had trouble keeping busy and active? | | | 620 | |
| 16. Does it take you longer to do the things you usually do? | | | .658 | |
| 17. Did you get the impression that you are doing things right (broadly speaking)? | | | .754 | |
| 18. Have you been satisfied with the way you do things? | | | .738 | |
| 19. Have you felt you have a useful role in life? | | | .469 | |
| 20. Do you feel able to make decisions? | | | .660 | |
| 21. Do you enjoy your normal activities every day? | | | .647 | |
| 22. You think you're a worthless person? | | | | .817 |
| 23. Do you live life totally without hope? (the last few weeks) | | | | .846 |
| 24. Do you feel that life is not worth living? | | | | .888 |
| 25. Have you thought about the possibility of "taking your own life"? | | | | .894 |
| 26. Have you noticed that sometimes you "can't do anything" because your nerves are very upset? | | | | .884 |
| 27. Do you wish you were dead and away from everything? | | | | .921 |
| 28. Have you noticed that the idea of taking one's life repeatedly comes to mind? | | | | .862 |
| | Correlations | | | |
| F1. Somatic symptoms | - | | | |
| F2. Anxiety/insomnia | .75 | - | | |
| F3. Social dysfunction | .62 | .63 | - | |
| F4. Severe depression | .55 | .63 | .64 | - |

In Table 5, the invariance of the four correlated factors from the CFA (confirmatory factor analysis) is shown. We found strict invariance, it is to be noted that the factorial loads are similar in the group of participants with diagnosis of psychological disorder and non-psychological cases, as well as in the group according to level of studies. However, according to gender (men and women), invariance is evidenced, where the configural model (baseline) presents adequate fit indices X2 (gl) = 595.11 (688), CFI = 0.90, RMSEA = 0.042, with reference to the model metric (weak invariance), scalar (strong invariance) and strict invariance.

The factor loadings between men and women are equal. There are no statistically significant differences (p> .05) and (ΔCFI ≤ .01), (ΔRMSEA ≤ .015) when comparing with the base model (configural), the values found are below the cut-off points established with respect to the metric invariance.

Finally, the strict invariance is observed (restrictions on factor loads, intercepts and residuals) that indicates variance in the group of men and women, with statistically significant differences (p <.05) and (ΔCFI ≤ .01), the values found were (p = 0.001), (ΔCFI ≤ .023), to verify these differences, the Student's t test and effect sizes were used, using the *d* of Cohen (1992), the referential values are, d = 0.20 (small), d = 0.50 (medium) and d = 0.80 (large), finding statistically significant differences and median TE in two factors, somatic symptoms (p <0.01; d = 41) and anxiety / insomnia (p <0.01; d = 35), the other two factors (social dysfunction and severe depression) did not show differences.

**Table 5**

*Measurement invariance for the GHQ-28 four-factor model, according to gender, diagnosis and latest studies performed*

| | Invariance | X2 (gl) | CFI | RMSEA | Δ x2 (Δ gl) | ΔCFI | ΔRMSEA |
|---|---|---|---|---|---|---|---|
| Gender | Configural | 595.11 (688) | .901 | .042 | | | |
| | Weak | 717.17 (712) | .924 | .036 | 8.22 (24) | .023 | .006 |
| | Strong | 728.72 (736) | .925 | .035 | 6.23 (24) | .001 | .001 |
| | Strict | 857.46 (740) | .902 | .040 | 13.36 (4) *** | .023 | .005 |
| Diagnosis | Configural | 1118.1 (688) | .735 | .045 | | | |
| | Weak | 1255.4 (712) | .710 | .046 | 28.13 (24) | .025 | .001 |
| | Strong | 1321.9 (736) | .701 | .046 | 19.35 (24) | .009 | .000 |
| | Strict | 4955.3 (740) | .000 | 113. | | .701 | .067 |
| Latest studies | Configural | 967.79 (1376) | .896 | .042 | | | |
| | Weak | 1436.91 (1448) | .914 | .037 | 21.99 (72) | .019 | .005 |
| | Strong | 1481.78 (1520) | .913 | .037 | 18.34 (72) | .001 | .001 |
| | Strict | 1604.95 (1532) | .905 | .038 | 8.45 (12) | .009 | 002 |

*Note:*** p <0.001; X2: Chi square; gl: degrees of freedom; ΔX2: Difference between the Chi square values; Δgl: Difference between degrees of freedom; CFI: comparative adjustment index; RMSEA: mean square root of the approximation error; ΔCFI: Difference between the comparative fit indices.
